# Assessing Global Neurosurgical Disease Burden Related To Climate Change: A Scoping Review

**DOI:** 10.1101/2025.10.04.25337272

**Authors:** Matthew Schaefer, Riya M. Dange, Caitlyn Smith, Daniela A. Perez-Chadid, Patricia Fuentes, Nathan A. Shlobin, Gail Rosseau

## Abstract

**Introduction:** Climate change is increasingly recognized as a significant threat to global health. Environmental exposures such as air pollution, temperature fluctuations, and extreme weather events may influence the presentation and outcomes of neurosurgical diseases. However, there is limited synthesis of the literature on this intersection.

**Objective:** To systematically map the global literature evaluating the relationship between planetary health and neurosurgical conditions, with a focus on exposure types, disease outcomes, and geographic and economic disparities in research output.

**Methods:** We conducted a scoping review following PRISMA-ScR guidelines. PubMed, Embase, and Scopus were searched from 1995 to 2024 for English-language, full-text studies reporting primary data on environmental exposures and neurosurgical disease. Studies were categorized by exposure type, outcome, study design, WHO region, and World Bank income classification.

**Results:** Of 15,658 records screened, 203 studies met inclusion criteria. Most were retrospective (24%, n = 49), prospective (17%, n = 34), or time series analyses (15%, n = 30). Studies originated primarily from high-income countries (62%, n = 126), with fewer from upper-middle-income (31%, n = 63), lower-middle-income (1%, n = 3), and low-income countries (<1%, n = 1). Western Pacific (40%, n = 81) and European (29%, n = 59) regions dominated the research. The most studied exposures were neurotoxic pollutants (30%, n = 61), PM2.5 (28%, n = 57), and temperature fluctuations (10%, n = 21). Ischemic stroke (31%, n = 63) and hemorrhagic stroke (29%, n = 58) were the most common outcomes, with notable links to PM2.5 and heat exposure. Pediatric diseases, epilepsy, and neuroinfectious conditions were underrepresented, as was research on surgical environmental impact.

**Conclusion:** There is increasing evidence linking environmental exposures to neurosurgical disease, particularly stroke. However, global disparities in research persist. Expanding research capacity and integrating planetary health principles into neurosurgery are essential for addressing emerging challenges in neurological health worldwide.

## Introduction

In 2022, the American Medical Association (AMA) declared climate change a “public health crisis that threatens the health and well-being of all individuals”^1^. The AMA’s subsequent advocacy for greenhouse gas reduction policies and clean energy solutions underscores the ties between human health and planetary health. Climate change, the amalgamation of long-term shifts in temperature and weather patterns on Earth, is considered a “fundamental threat to human health” by the World Health Organization (WHO)^2^. Currently, 3.6 billion people live in areas deemed highly susceptible to climate change. By 2030, an estimated 250,000 additional deaths will occur annually due to climate-related causes, and climate change is projected to cost the U.S. between $2 billion and $4 billion annually. While climate change-driven heat waves, floods, tropical storms, and hurricanes pose overt hazards to human safety, the rising frequency of infections from zoonotic, food-borne, water-borne, and vector-borne pathogens is also an important, insidious threat. These environmental risk factors disproportionately impact people of certain geographic regions and lower-income communities. It is necessary to combat climate change so human health can flourish.^2^

Climate change may affect neurological diseases that require surgical and endovascular intervention. The most prevalent neurosurgical conditions that have been found to have an association with environmental exposures are hemorrhagic and ischemic stroke, seizure, central nervous system (CNS) cancer, and brain abscesses, which have all been found to be increased by such as neurotoxic pollutants [ex: PM 2.5, PM 10, sulfate (SO2−4), nitrous oxide (NO), and nitrogen dioxide (NO2)], neuroinfectious diseases, extreme weather events, and temperature fluctuations^3,4^. Understanding the role of climate change in neurosurgery is important, especially when viewed through the lens of LMICs, which have higher burdens of neurosurgical disease, as well as higher risk of increasing burden of disease with climate change.

There is a paucity of literature on climate change with respect to neurosurgery. Our study is a scoping review of the current literature regarding planetary health and neurosurgical conditions. The primary aim is to investigate the impact of climate change on neurosurgical diseases through a planetary health lens and on a global scale. The second aim is to understand how LMIC and HIC (high-income countries) differ in their burden of disease and published literature on neurosurgical conditions potentially related to environmental factors. The third aim is to elucidate how neurosurgical care is contributing to environmental changes and provide guidance for best practices, based on peer-reviewed research. This is the first study to comprehensively investigate how neurosurgical conditions are changing over time in their relationship to environmental exposures.

## Methods

A scoping review was performed following the guidelines established by the Preferred Reporting Items for Systematic Reviews and Meta-Analyses Extension for Scoping Reviews to investigate the impact of climate change in neurosurgery as well as the intersection between planetary health and research related to neurosurgical conditions^5^. A trained medical library-science specialist supported our team in building the search structure and process. PubMed, Embase (Elsevier), and Scopus (Elsevier) were searched on June 27, 2024. There were no restrictions on language, date, or article type. A comprehensive search was conducted, as seen in Supplemental Figure 1. The keywords used in the search strategy focused on neurosurgery (including the terms “neurosurgery”, “brain injuries”, “spinal cord injuries”, and “brain tumors”) and climate change (covering concepts including “global warming”, “wildfires”, “extreme weather”, and “carbon footprint”). Additionally, terms related to planetary health and environmental impacts (such as “environmental degradation”, “air pollution”, and “vector-borne disease”) were included to explore the intersection between climate change and neurosurgical practices.

Inclusion criteria included articles with full-text availability, written in or translated into the English language, and pertaining to neurosurgery and planetary health. The included articles were focused on neurosurgical pathology with potential for a need for neurosurgical intervention. Exclusion criteria included articles not published or available in English, without full data available, not pertaining to neurosurgery, and not discussing planetary health. Also excluded from the criteria are conference abstracts, case reports, narrative reviews, scoping reviews, systematic reviews, and meta-analyses.

The scoping review was conducted through the open-access website Rayyan. After the initial articles were determined, duplicates were excluded. Three individuals then independently conducted a blinded screening of titles and abstracts. Any conflicts that arose where the 3 blinded researchers did not agree to include or exclude the study, a fourth researcher determined whether the study would be included or excluded. The included studies then went through a full-text review where two participants independently decided to include or exclude the study based on prespecified inclusion and exclusion criteria. This thorough approach ensured a consistent and unbiased evaluation of the relevant literature. Lastly, we hand-searched literature for important research that may have been missed by our database search. This included reviewing the reference lists of relevant review articles and included studies to identify additional eligible articles.

### Study Categorization and Analyses

Included studies were categorized by design, year of publication, and the World Health Organization region of their institution of origin. Income classifications were assigned on the country level by the World Bank Income Classification index for the financial year 2025^6^.

Independent reviewers coded the studies binarily for the discussion of eight exposure topics and seven neurosurgical disease topics, pre-selected based on clinical interest. The exposure topics included: PM2.5, neurotoxic pollutants, neuroinfectious disease, extreme weather events, temperature fluctuations, pesticides, carbon emissions, and noise pollution. The disease topics included: ischemic stroke, hemorrhagic stroke, seizure or epilepsy, pediatric or developmental disease, central nervous system cancer, intracranial hemorrhage, and brain abscess. Descriptive analyses were conducted in R for all classifications of interest.

### Ethical Considerations

No institutional review board (IRB) approval was required for this study, as all analyses were secondary and conducted on anonymized, published data.

## Results

### Study Selection and Publication Trends

The study selection process is detailed in Figure 1, which presents a PRISMA flow diagram outlining the inclusion and exclusion criteria applied. There were a total of 15,658 total articles identified through PubMed, Scopus, and Embase. The initial search terms can be found in Supplementary Table 1. A total of 5,264 articles were excluded for duplication, leaving 10,394 articles screened by title and abstract. 10,101 articles were excluded based on title and abstract, resulting in a total of 293 articles eligible for full-text screening. Out of the 293 full-text articles screened, 90 articles were excluded based on exclusion criteria or at least 2 researchers not having access to the full text. Lastly, the manual search retrieved 8 articles. This resulted in 211 articles being included in the review (Figure 1). Figure 2 provides a visual representation of the distribution of studies across key variables.

### Study Characteristics and Geographic Regions

The 211 included studies were published between 1995 and 2024, with the greatest number of publications in 2023 (Table 1). The most common study designs were retrospective cohort studies (*n* = 49, 24%), followed by prospective cohort studies (*n* = 34, 17%), then time series analyses (*n* = 30, 15%) and case-crossover studies (*n* = 30, 15%). We set out to investigate how published studies regarding planetary health and neurosurgical diseases differed between WHO regions, as well as how they differed when analyzing World Bank Income Classification groups. The highest representation was from the Western Pacific (40%) and Europe (29%), while Africa had the lowest number of studies (1%) (Table 1). In terms of World Bank Income Classification, most studies were conducted in high-income countries (HICs) (62%), followed by upper-middle-income countries (UMICs) (31%) (Table 1). Lower-middle-income (LMICs) and low-income countries (LICs) were significantly underrepresented, accounting for only 1% and <1% of studies (Table 1).

**Table 1.**
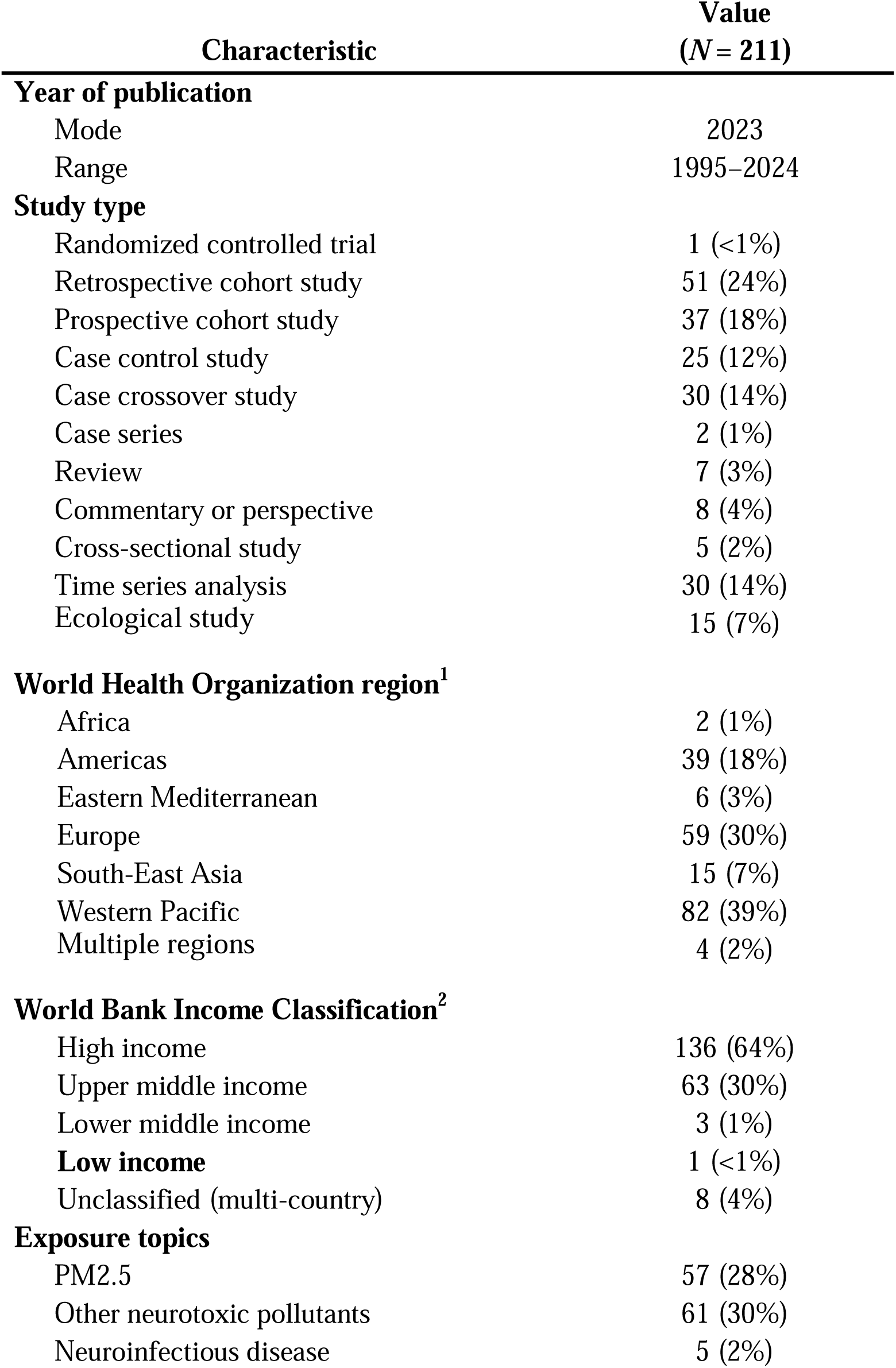
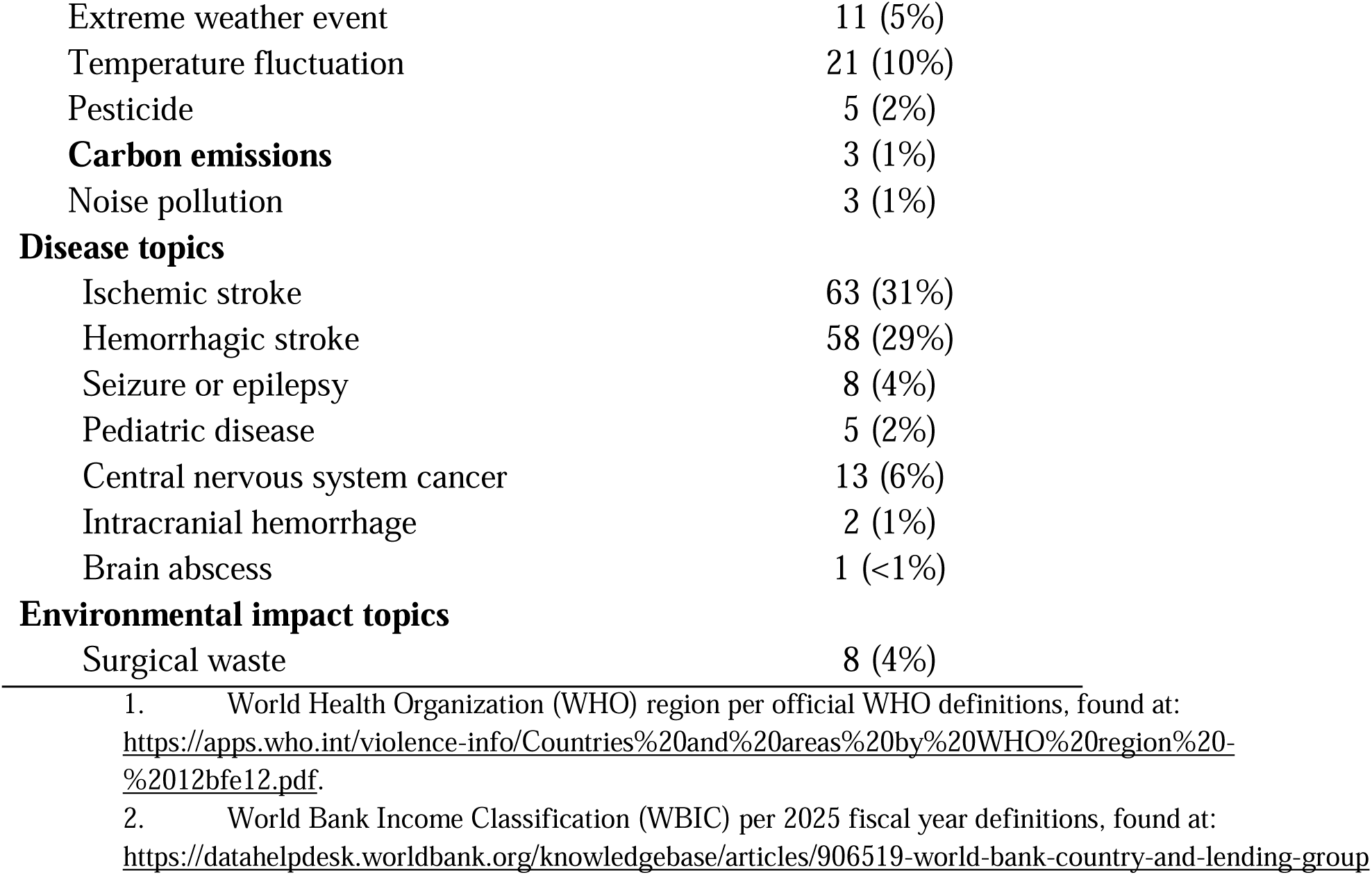
Characteristics of included studies.

| Characteristic | Value<br>(*N* = 211) |
| --- | --- |
| **Year of publication** |  |
| Mode | 2023 |
| Range | 1995–2024 |
| **Study type** |  |
| Randomized controlled trial | 1 (<1%) |
| Retrospective cohort study | 51 (24%) |
| Prospective cohort study | 37 (18%) |
| Case control study | 25 (12%) |
| Case crossover study | 30 (14%) |
| Case series | 2 (1%) |
| Review | 7 (3%) |
| Commentary or perspective | 8 (4%) |
| Cross-sectional study | 5 (2%) |
| Time series analysis | 30 (14%) |
| Ecological study | 15 (7%) |
| **World Health Organization region<sup>1</sup>** |  |
| Africa | 2 (1%) |
| Americas | 39 (18%) |
| Eastern Mediterranean | 6 (3%) |
| Europe | 59 (30%) |
| South-East Asia | 15 (7%) |
| Western Pacific | 82 (39%) |
| Multiple regions | 4 (2%) |
| **World Bank Income Classification<sup>2</sup>** |  |
| High income | 136 (64%) |
| Upper middle income | 63 (30%) |
| Lower middle income | 3 (1%) |
| **Low income** | 1 (<1%) |
| Unclassified (multi-country) | 8 (4%) |
| **Exposure topics** |  |
| PM2.5 | 57 (28%) |
| Other neurotoxic pollutants | 61 (30%) |
| Neuroinfectious disease | 5 (2%) |
| Extreme weather event | 11 (5%) |
| Temperature fluctuation | 21 (10%) |
| Pesticide | 5 (2%) |
| **Carbon emissions** | 3 (1%) |
| Noise pollution | 3 (1%) |
| **Disease topics** |  |
| Ischemic stroke | 63 (31%) |
| Hemorrhagic stroke | 58 (29%) |
| Seizure or epilepsy | 8 (4%) |
| Pediatric disease | 5 (2%) |
| Central nervous system cancer | 13 (6%) |
| Intracranial hemorrhage | 2 (1%) |
| Brain abscess | 1 (<1%) |
| **Environmental impact topics** |  |
| Surgical waste | 8 (4%) |

- 1. World Health Organization (WHO) region per official WHO definitions, found at: https://apps.who.int/violence-info/Countries%20and%20areas%20by%20WHO%20region%20-%2012bfe12.pdf. 2. World Bank Income Classification (WBIC) per 2025 fiscal year definitions, found at: https://datahelpdesk.worldbank.org/knowledgebase/articles/906519-world-bank-country-and-lending-group

### Exposure and Disease Topics

The most frequently studied exposure topics included neurotoxic pollutants such as PM 10, NO2, sulfate trioxide (SO3) (30%), PM2.5 air pollution (28%), and temperature fluctuations (10%) (Table 1). Fewer studies focused on extreme weather events (5%), neuroinfectious disease (2%), pesticide exposure (2%), carbon emissions (1%), and noise pollution (1%) (Table 1). In terms of disease focus, ischemic stroke (31%) and hemorrhagic stroke (29%) were the most common conditions examined, while fewer studies addressed seizure disorders (4%), pediatric neurological diseases (2%), and brain abscesses (<1%) (Table 1). The heatmap showed that the greatest environmental exposure and neurological disease relationship is between PM 2.5 and stroke (hemorrhagic and ischemic), followed by other neurotoxic pollutants and stroke (hemorrhagic and ischemic), and then temperature fluctuations and stroke (hemorrhagic and ischemic) (Table 3). A substantial number of studies discussed the association of PM 2.5 and CNS tumors, as well as other neurotoxic pollutants and CNS tumors (Table 3).

### Associations between Environmental Exposures and Neurological Diseases

Utilizing the heatmaps, the studies reviewed identified associations between a majority of the environmental exposures (PM 2.5, other neurotoxic pollutants, temperature fluctuations/ extreme heat, pesticides, extreme weather, noise pollution, and carbon emission) and neurosurgical diseases (Table 3 and Table 4).

PM 2.5 exposure was the most frequently studied environmental factor. Forty-two of the 70 studies investigating the association between PM 2.5 and hemorrhagic stroke and 42 of the 67 studies investigating the association between PM 2.5 and hemorrhagic stroke reported a statistical significance, significance, or association (Table 4). Multiple studies found that exposure to PM 2.5 was associated with an increased risk of both hemorrhagic and ischemic stroke. One cohort study in an East Asian population observed that long-term PM2.5 exposure correlated with elevated stroke risk and all-cause mortality, even after adjusting for urban density and comorbid conditions^7^. Studies conducted in South America and China reported that increases in [PM 2.5] and ambient air pollution were positively associated with hospital admissions for ischemic and hemorrhagic stroke, as well as increased stroke mortality^8–10^. Not all studies, however, found an association between [PM 2.5] and stroke. Cui and colleagues reported no significant association between short-term exposure to gaseous air pollutants (NO2, SO2, CO, O3) and ischemic stroke when controlling for meteorological variables, illustrating some heterogeneity in findings^11^.

Pesticide exposure was found to have statistical significance or association with hemorrhagic stroke, seizure/epilepsy, and CNS cancer in a majority of the studies investigating pesticide exposure (Table 3 and Table 4). Carod-Artal and colleagues described a statistically significant association between pesticide exposure and the development of meningioma, whereas statistical significance with glioma was not found. The potential link between neurotoxic agricultural chemicals and tumorigenesis requires further study^12^.

Seventy-nine studies focused on the relationship between temperature fluctuation/ extreme weather events and neurosurgical disease. These studies reported that daily stroke was positively associated with sudden drops in temperature, surging airborne irritants after dust storms, and extreme heat waves^13,14^. While not all results reached statistical significance, the overall evidence supports a growing concern that thermal variability and extreme climate events may exacerbate or trigger neurosurgical disease presentations, particularly strokes and hemorrhagic events.

Eight studies examined the impact that neurosurgery creates on environmental and planetary health^15–22^. Rigante et. al. found that, on average, each neurosurgery procedure was associated with €515 in unused disposal products that had to be disposed of after the procedure ^20^. Emerging research on sustainability in neurosurgery reveals consistent patterns of excessive resource use, high waste generation, and underutilized opportunities for environmental stewardship ^20^. Across a range of neurosurgical procedures, significant proportions of disposable surgical supplies are routinely opened but left unused, leading to both financial inefficiencies and substantial material waste. Neurosurgical operations also contribute substantially to hospital waste and carbon emissions, with energy consumption, single-use items, and sterilization processes identified as major contributors. Despite the clear environmental impact, the implementation of sustainable practices remains limited. These findings underscore an urgent need for neurosurgery to integrate environmental responsibility into clinical and operational workflows, aligning the specialty with broader planetary health goals.

## Discussion

### 1. Purpose of study and key findings

The purpose of this study was to identify how the changing climate is affecting neurosurgical diseases and care. We summarized the literature to better understand the relationship between environmental changes and neurosurgical diseases and care, while focusing on how low, middle, and high-income countries differ in their research output and approach to care. We identified ways neurosurgical care is contributing to environmental changes. Our study highlights that 1) abiotic environmental exposures such PM 2.5 and extreme weather are negatively associated with certain neurosurgical diseases such as stroke and seizure, 2) high income countries have led the research pertaining to neurosurgery care and planetary health, although low income countries have a greater burden of neurosurgical diseases, and 3) neurosurgical care exerts tangible impacts on planetary health, particularly in the operating room, that can be reduced.

Although an estimated 17.6 million patients in low- and middle-income countries (LMICs) require neurosurgical care—compared to 4.3 million in high-income countries (HICs)—less than 1% of published research on planetary health-related neurosurgical conditions originates from LMICs. Informing the neurosurgical community of such associations between planetary health and neurosurgery, addressing root causes of environmentally related neurosurgical conditions and implementing ways to reduce the negative impact on planetary health of our discipline can lead to a lower burden of neurosurgical diseases.

### 2. The Current State of Planetary Health in Neurosurgery

#### PM 2.5, other pollutants, and stroke

Through this scoping review, we found that the majority of research for neurosurgical disease exacerbated by environmental exposure concerns PM 2.5, other neurotoxic pollutants (black carbon (BC), organic matter (OM), SO2−4), and stroke. A major concern for neurological health is PM2.5 accumulation in the brain. PM2.5 is a neurotoxin that easily crosses the blood-brain barrier and activates the innate immune responses in the astrocytes, microglia, and neurons, thus exerting neurotoxicity^23,24^. Cai et. al. performed an exposure assessment characterizing the concentration of PM2.5 associated with increased risk of stroke, and the components of PM2.5 that increase the risk of stroke. They found that a [PM2.5] of 15.14 ug/m^3 is statistically significantly associated with an increased risk of stroke, with an odds ratio of 1.131 (CI of 1.118–1.157). In 2024, the Environmental Protection Agency (EPA) set the air concentration standard of 9.0 ug/m^3 for PM2.5. Although this concentration is below the amount found to be associated with increased cause of stroke, many areas of the world have [PM2.5] greater than 15.14 ug/m^3.^25^ The highest concentrations of PM2.5 are found in Asia and Africa, with most regions in these continents reporting levels between 27–85 µg/m³. In contrast, the United States remains below the 15.14 µg/m³ threshold associated with an increased risk of stroke. However, many low- and middle-income countries (LMICs), and even some high-income countries (HICs), exceed this threshold, placing their populations at greater risk.² Increased incidence of severe storms leads to an increased amount of pesticide runoff into water sources According to Zhang and colleagues, 66% of the countries in the world are experiencing wetter environments compared to the past.^26^ Studies have identified an association between pesticide exposure and increased odds of epilepsy. For example, a study by Alarcon and colleagues discovered that pesticide exposure is associated with up to a 3.4-fold increased incidence of epilepsy, more commonly amongst the farming workforce^27^. In our study, only 5 studies (2.3%) that investigated the relationship between pesticide use and the incidence of epilepsy met the inclusion criteria. This relationship should be further studied to enable the implementation of effective protection measures. Utilizing a pesticide reduction policy significantly reduces the annual discharge load of pesticides within a watershed area^28^. Contaminated drinking water has been shown to negatively affect a child’s neurological development by causing arsenic, nitrates, and lead to build up in a child’s developing nervous system. Congenital infections such as toxoplasmosis and Zika virus are increasingly influenced by climate change and can result in serious neurological complications, including seizures, cognitive impairment and hydrocephalus. Rising temperatures and rainfall have been linked to increased prevalence of both *T. gondii* and Zika, particularly in low-resource, climate-vulnerable regions^29–32^. These infections not only burden affected children and families but also strain healthcare systems, especially where neurosurgical access is limited. Recent data estimate that there are approximately 72,967 neurosurgeons worldwide, with a global median density of only 0.44 per 100,000 people—far below the target of 1 per 100,000 in most LMICs ^33^. The growing impact of climate-sensitive congenital infections highlights a pressing planetary health challenge and underscores the urgent need to expand neurosurgical capacity for the 1.2 billion children living in LMICs..^34^

### 3. Planetary Health Inside and Outside of the Operating Room

Carbon dioxide (CO2) is a well-known greenhouse gas emission. Surgical departments are among the most resource-intensive areas of hospitals, contributing between 21% and 30% of the entire hospital’s CO2 waste, and consuming three to six times more energy than other units. A substantial proportion of hospital solid and regulated medical waste also originates from operating rooms, with estimates indicating that it accounts for 50 to 70% of the total waste stream ^35^. Within neurosurgery, single-use equipment and disposables generate on average 8.9 kg of waste per case, which converts to 24.5 CO2 equivalents (CO2e)^18^. Single-institution data have reported that the annual waste contributions of a single neurosurgical facility can reach 11,584. kg/year and a carbon footprint of 31,869 kg CO2e.^18^ These values are highly dependent on institutional size and case volume; the UCSF analysis of 58 cases projected an annual waste cost of $2.9 million for its neurosurgical service, reflecting $653 of unused disposable supplies per case (13.1 % of supply cost) ^17^. While the precise share of global carbon emissions attributable to neurosurgery is not reported, health care overall contributes about 8 % of national greenhouse gas output, and anesthesiology alone accounts for about 4.5 % of global GHG emissions ^17,36^. To address these impacts, a Japanese “Green Hospitals” framework proposes eight stewardship principles—rethink, refuse, reduce, reuse, recycle, research, renovation, and revolution—guiding systematic waste and emission reductions, and its dissemination across hospital systems is advocated as a positive step toward lowering the environmental footprint of neurosurgical practice ^18^.

Choice of anesthetic technique represents another major modifiable factor. In spinal fusion procedures, general anesthesia with desflurane has reportedly been associated with emissions of roughly 22,700 g CO, compared with only 62 g for spinal anesthesia alone ^37^. The rising volume of spinal surgery warrants a closer look at the type of anesthetic gas used for surgical procedures and its broader impact on global carbon emissions. ^37^

Anesthesiology has therefore become a leading field in perioperative sustainability. Volatile agents such as desflurane and nitrous oxide contribute disproportionately to emissions, releasing up to 15–20 times the CO e per MAC-hour compared with sevoflurane or isoflurane ^38^. Current mitigation strategies follow a “reduce refine replace” model: employing low fresh gas flow (≤ 3 L min ¹), substituting high impact volatile agents with total intravenous or regional techniques, and phasing out desflurane and nitrous oxide ^38,39^. Departmental interventions, such as staff education, green team formation, and waste management protocols, have achieved reductions of 60% to 80% in CO e per case, making anesthetic stewardship a practical exemplar for neurosurgical sustainability initiatives ^39^.

### 4. Building Priorities for Planetary Health in Neurosurgery

To advance the understanding of the relationships between planetary health and neurosurgical conditions, researchers and public health officials will need to adopt region-specific strategies tailored to the unique challenges faced by HICs and LMICs. In HICs, efforts should focus on reducing environmental risk factors through stricter air quality regulations, climate-conscious healthcare policies, investments in green infrastructure and interdisciplinary collaborations between scientists, environmental health experts, and policymakers to facilitate the development of early warning systems and predictive models for climate-related neurological conditions, such as stroke and neuroinfectious diseases. In contrast, low-middle-income countries require targeted strategies that prioritize capacity building, equitable access to healthcare, and climate adaptation measures. Strengthening healthcare infrastructure, expanding neurosurgical services, and integrating planetary health considerations into public health programs are important next steps. Investment in surveillance systems to track environmental exposures and their neurological impacts can inform locally appropriate interventions. Global partnerships, technology transfer initiatives, and international funding mechanisms can support these efforts, ensuring that vulnerable populations in resource-limited settings receive adequate protection from the growing neurosurgical burden associated with planetary health challenges.

Hospitals can collaborate with neurosurgical and other surgical departments to host educational workshops on how neurosurgery care affects environmental and planetary health. Surgical team education, supply tracking, preference card optimization, adoption of reusable instruments, and integration of digital technologies have demonstrated the potential to reduce waste and emissions without compromising patient care. Neurosurgery conferences can provide resources for the neurosurgery community to learn about the relationship between neurosurgery and planetary health, as well as to discuss mitigation strategies that neurosurgeons can use to minimize their environmental footprint.

### 5. Limitations and Future Directions

This scoping review has several limitations. First, the inclusion of studies was limited to those available in selected databases, written or translated to English, and studies where researchers had access to full articles, which may have led to the omission of other relevant research papers. Second, while efforts were made to capture a comprehensive range of studies, variability in study design, geographic area assessed among individual studies, exposure assessment, and outcome definitions may have introduced heterogeneity among studies analyzed, limiting direct comparisons across studies. Third, the geographic distribution of research was skewed toward high-income countries, potentially underrepresenting the impact of planetary health factors on neurosurgical conditions in low and middle-income regions where environmental exposures and healthcare infrastructure differ significantly. Given the broad scope of planetary health and its intersections with neurosurgery, certain emerging risk factors or disease associations may not have been adequately covered.

Future systematic reviews and meta-analyses are warranted to further quantify the impact of planetary health on neurosurgical conditions. Using not only quantifiable data, but qualitative questionnaires and focus groups to better understand health care workers, government officials, and other stakeholders’ opinions for how to best address neurosurgical care and the changing environment. Planetary health is a global phenomenon that requires a global approach to help mitigate neurosurgical diseases that are exacerbated by environmental exposures and the changing climate.

### Conclusion

The burden of neurosurgical disease is related to our planetary health. As our environment changes, so do neurosurgical conditions that must be addressed. Collaboration among researchers, public health workers, government officials, and healthcare workers can lead to preventative measures that will decrease the incidence of neurosurgical disease, particularly stroke. This study shows research addressing planetary health and neurosurgical disease is increasing, especially for HICs, but LMICs have produced much less research, despite the disproportionate effect of climate change on their populations. Global health measures that address planetary health will likely decrease the global burden of neurological disease.

## Data Availability

All data produced in the present study are available upon reasonable request to the authors

## Acknowledgement

The authors thank Molly Beestrum, medical librarian at Northwestern University, for her invaluable assistance in developing and refining the literature search strategy.

The authors would like to disclose the use of Grammarly for editing purposes.

This article has not yet been peer-reviewed.

---

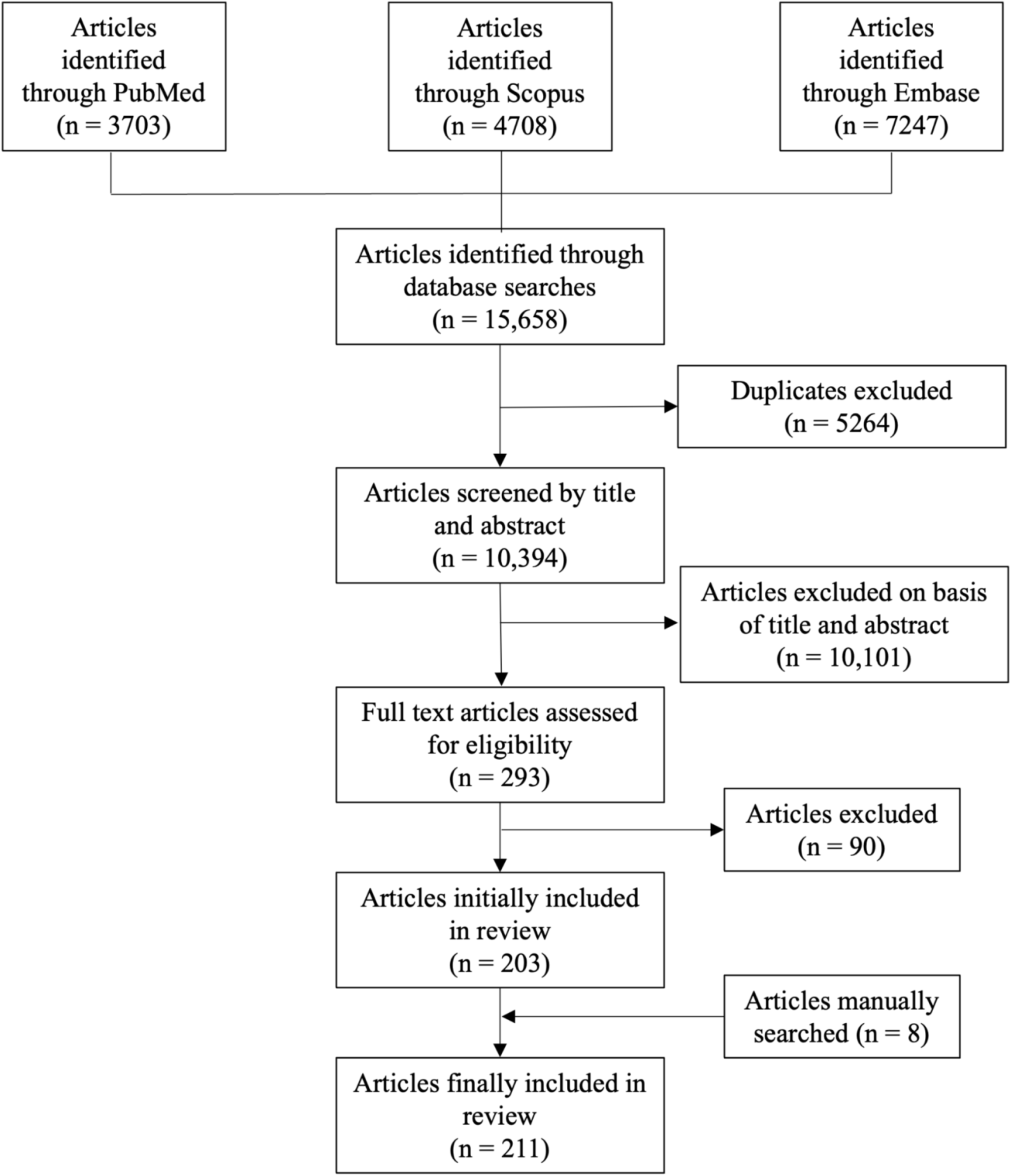

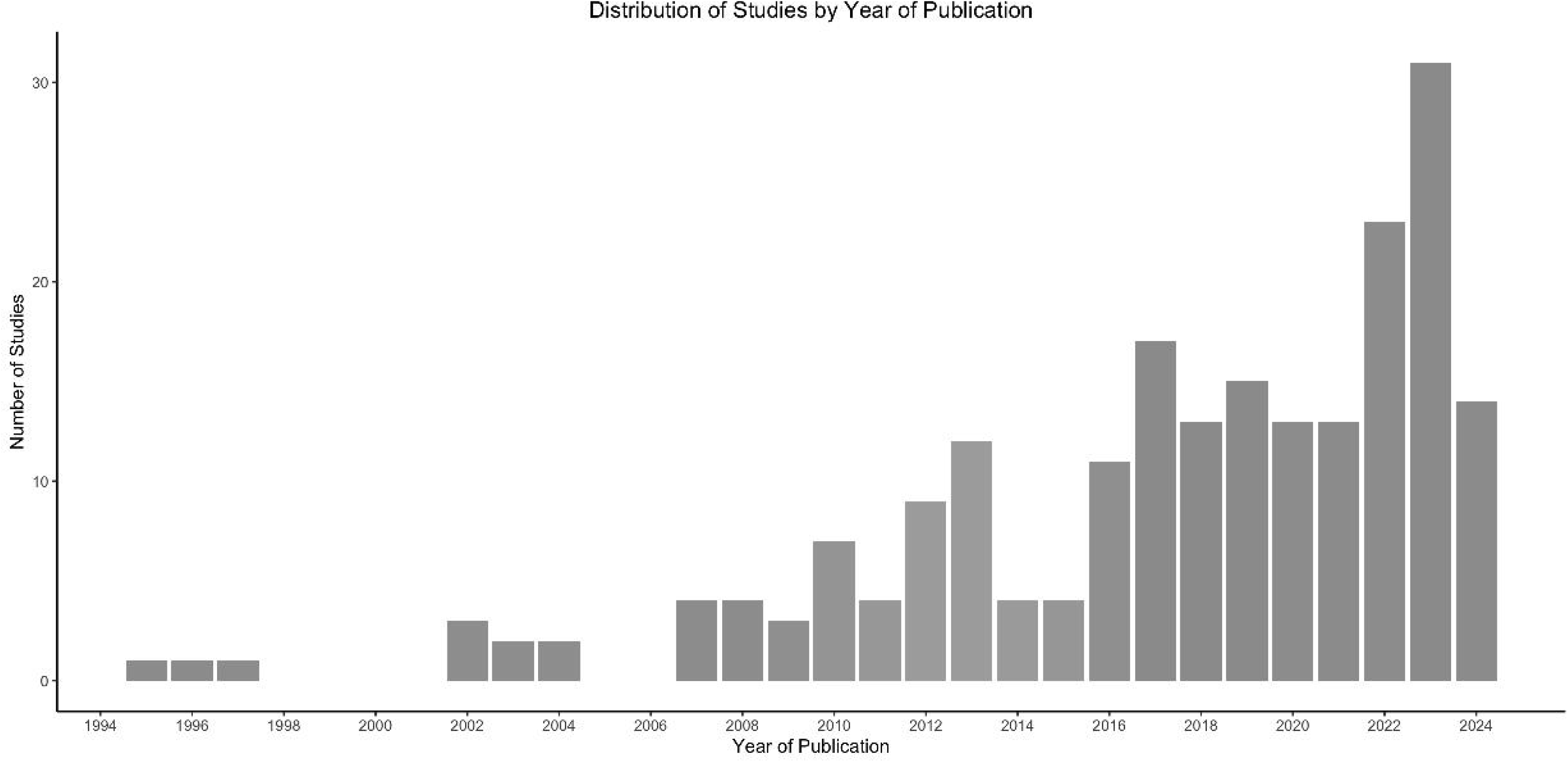

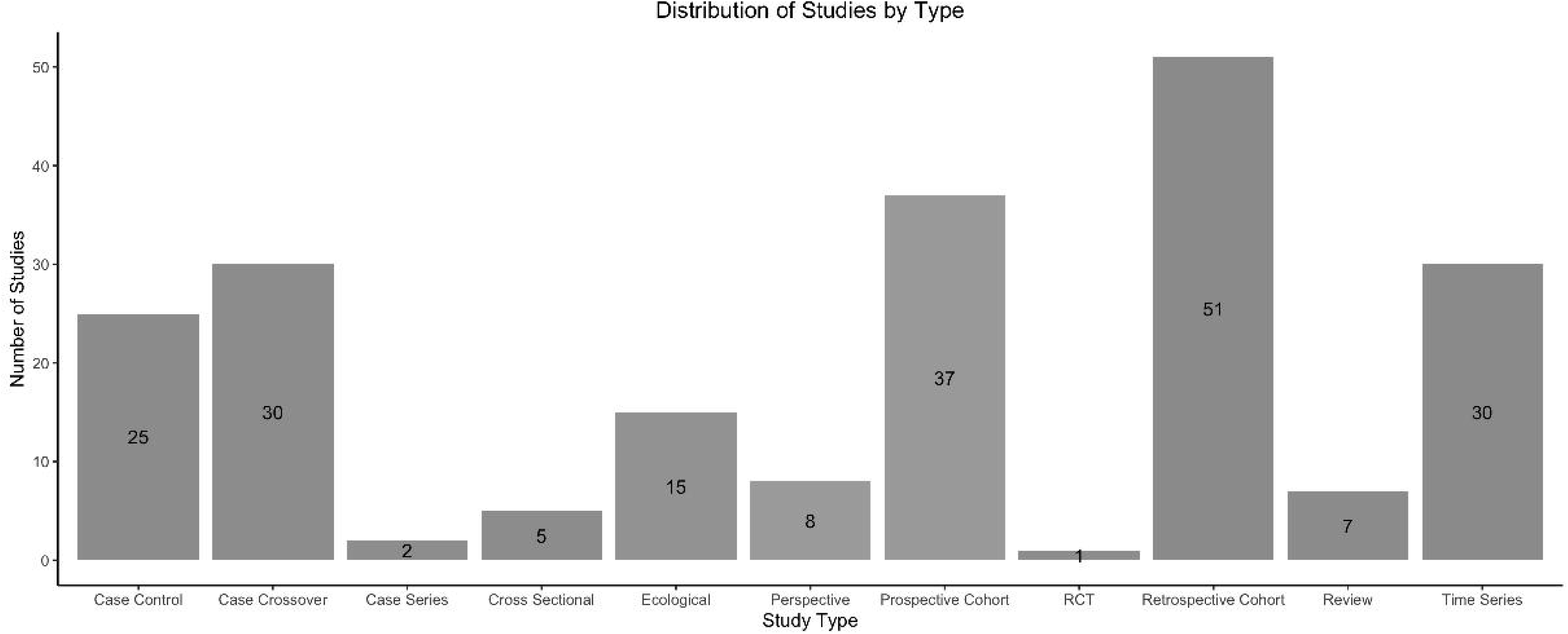

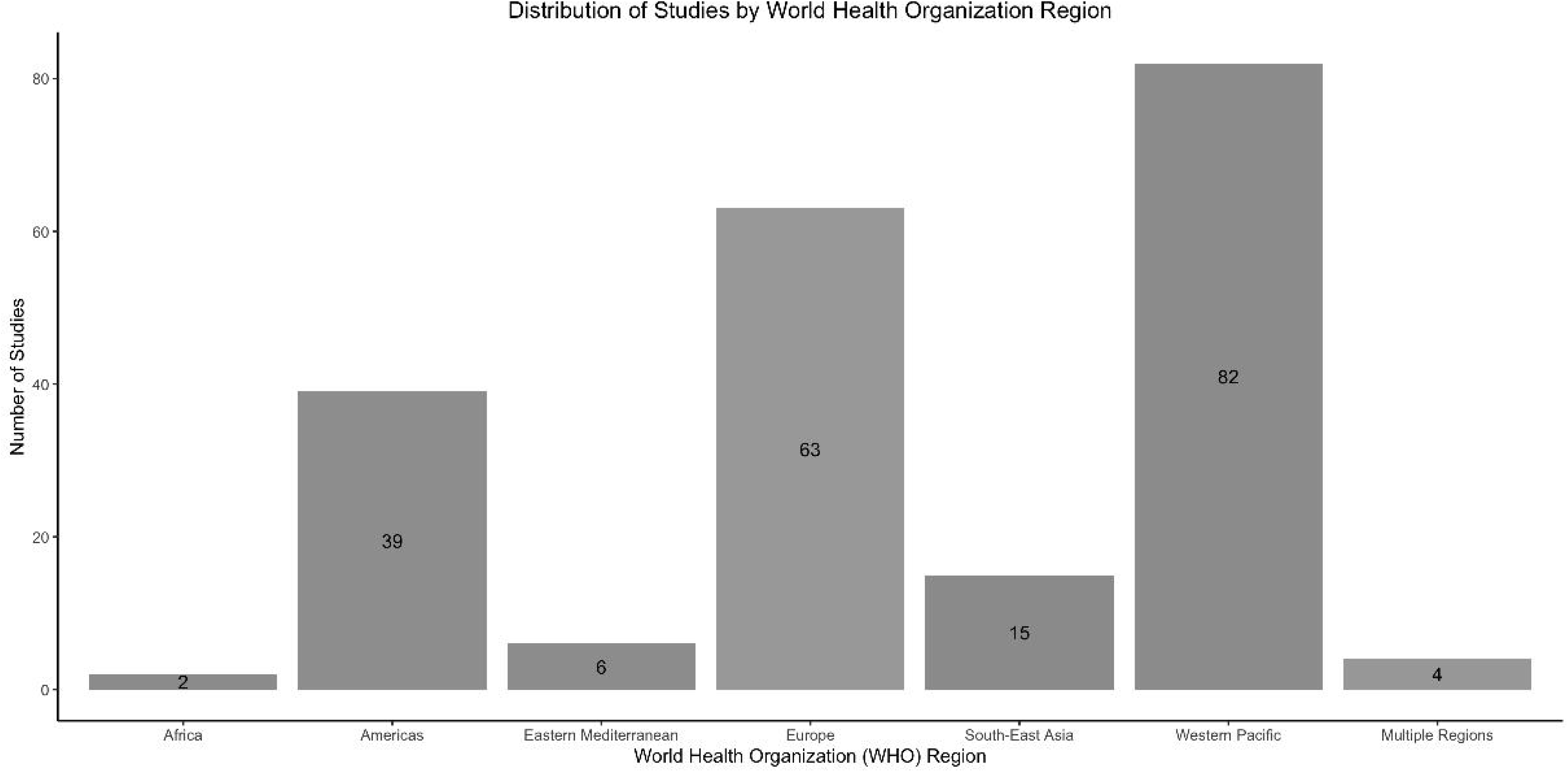

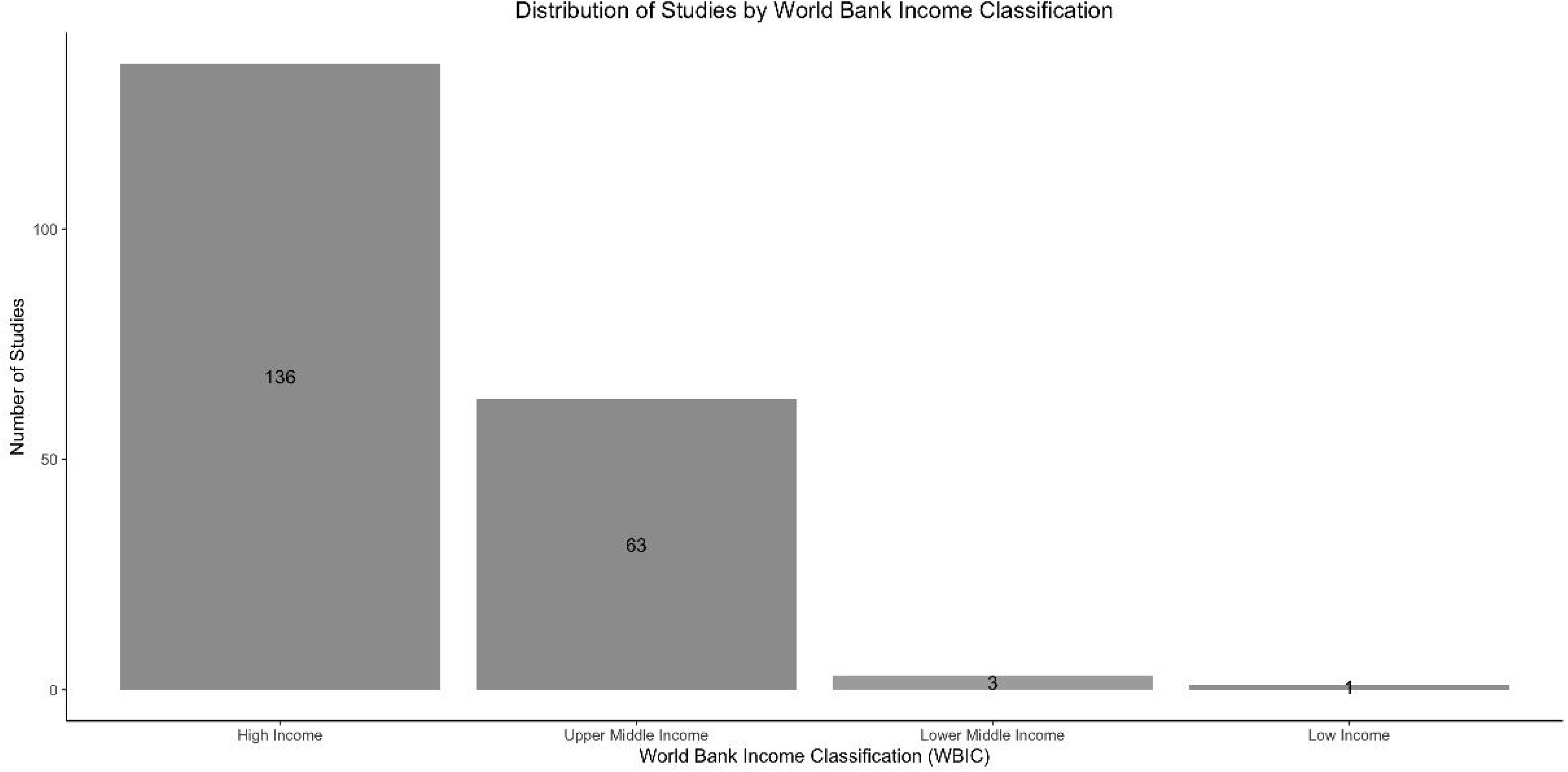

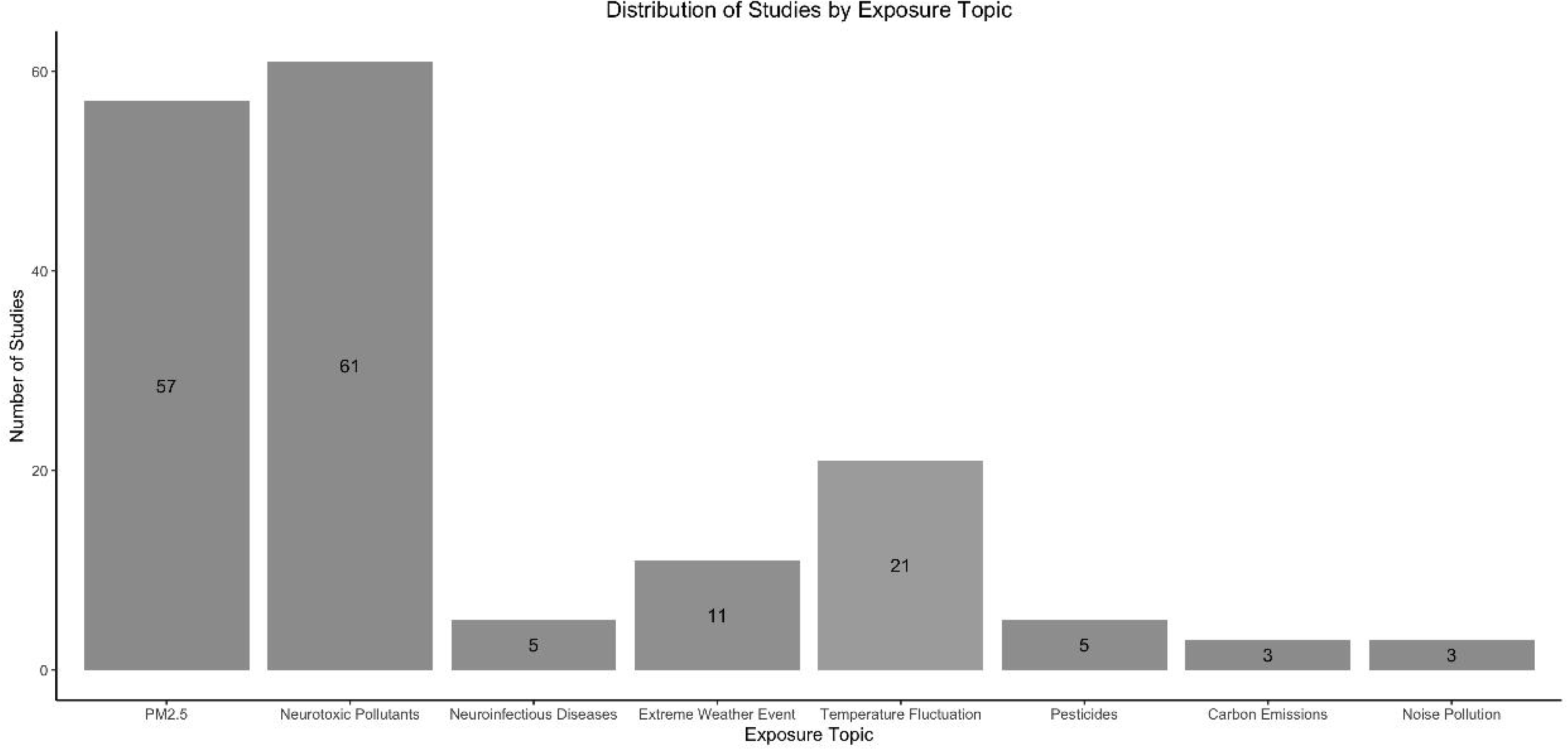

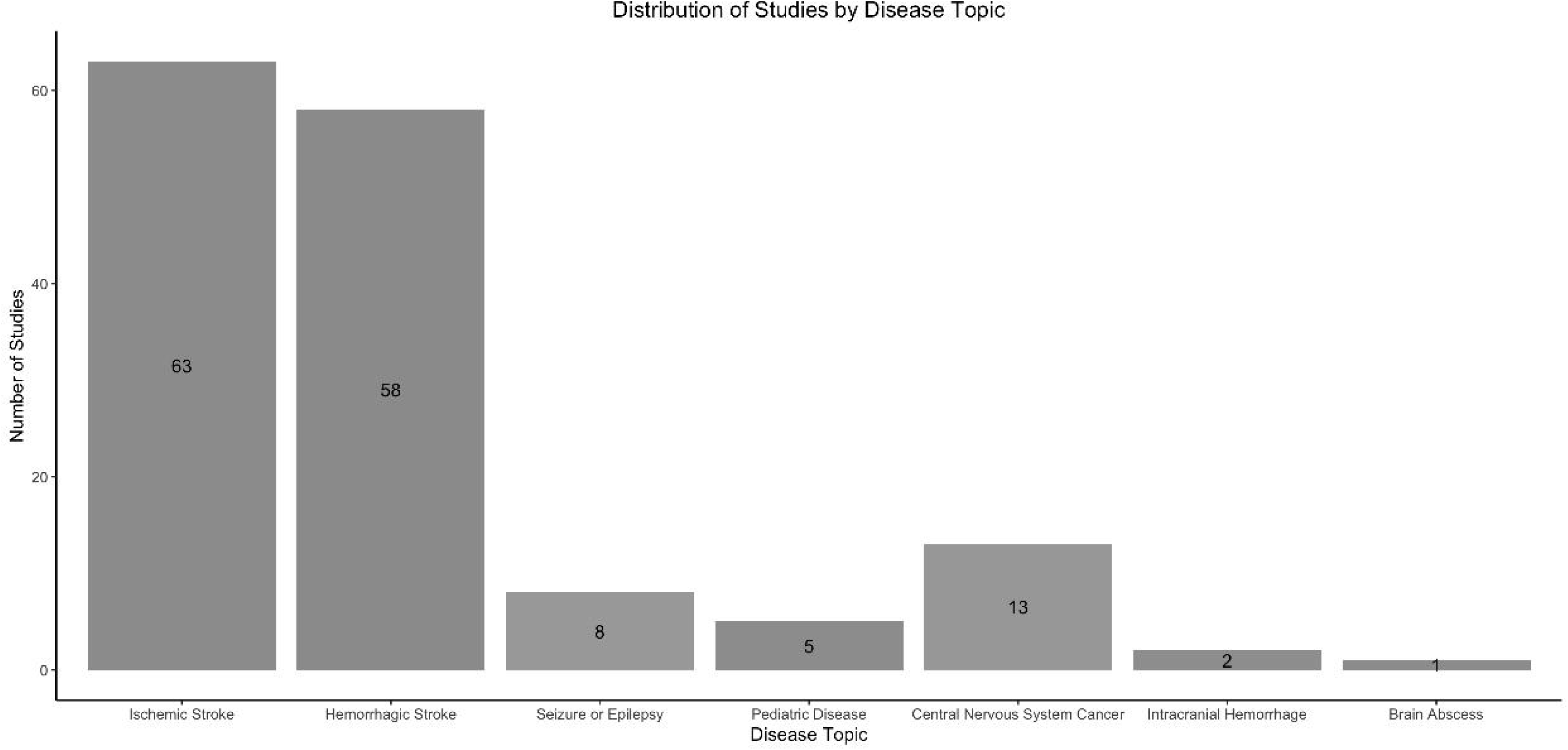

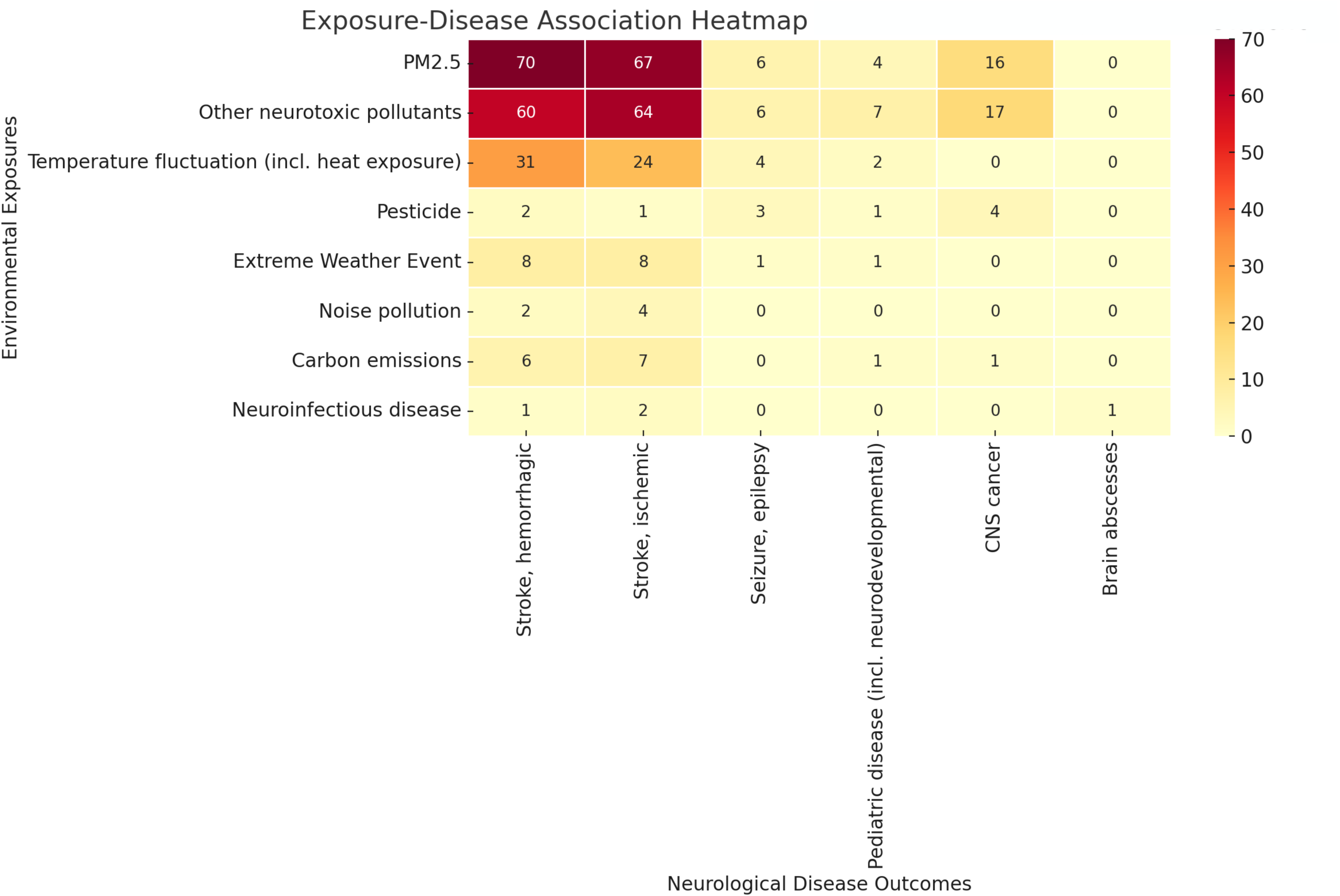

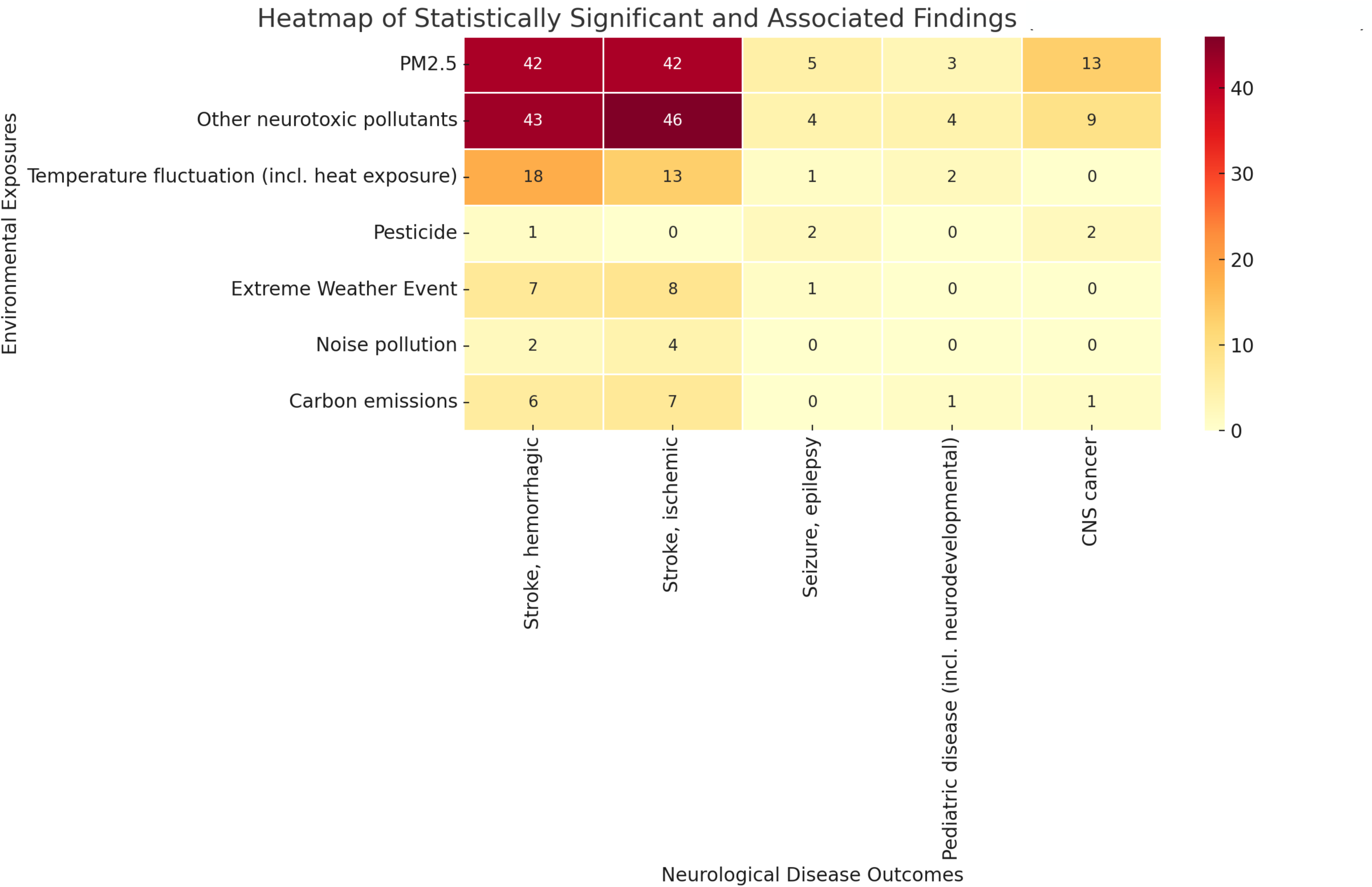

## References

1. D-135.966 Declaring Climate Change a Public Health Crisis | AMA. Accessed May 18, 2024. https://policysearch.ama-assn.org/policyfinder/detail/climate%20change?uri=%2FAMADoc%2Fdirectives.xml-D-135.966.xml

2. Climate change. Accessed May 18, 2024. https://www.who.int/news-room/fact-sheets/detail/climate-change-and-health

3. Kulick ER, Kaufman JD, Sack C. Ambient Air Pollution and Stroke: An Updated Review. Stroke. Published online March 2023. doi:10.1161/STROKEAHA.122.035498

4. Louis S, Carlson AK, Suresh A, et al. Impacts of Climate Change and Air Pollution on Neurologic Health, Disease, and Practice: A Scoping Review. Neurology. 2023;100(10):474–483. doi:10.1212/WNL.0000000000201630

5. Page MJ, McKenzie JE, Bossuyt PM, et al. The PRISMA 2020 statement: an updated guideline for reporting systematic reviews. BMJ. 2021;372:n71. doi:10.1136/bmj.n71

6. World Bank Country and Lending Groups – World Bank Data Help Desk. Accessed March 13, 2025. https://datahelpdesk.worldbank.org/knowledgebase/articles/906519-world-bank-country-and-lending-groups

7. Chen PC, Sung FC, Mou CH, et al. A cohort study evaluating the risk of stroke associated with long-term exposure to ambient fine particulate matter in Taiwan. Environ Health. 2022;21(1):43. doi:10.1186/s12940-022-00854-y

8. Leiva G MA, Santibañez DA, Ibarra E S, Matus C P, Seguel R. A five-year study of particulate matter (PM2.5) and cerebrovascular diseases. Environ Pollut Barking Essex 1987. 2013;181:1–6. doi:10.1016/j.envpol.2013.05.057

9. Chen R, Zhang Y, Yang C, Zhao Z, Xu X, Kan H. Acute effect of ambient air pollution on stroke mortality in the China air pollution and health effects study. Stroke. 2013;44(4):954–960. doi:10.1161/STROKEAHA.111.673442

10. Zhang R, Liu G, Jiang Y, et al. Acute Effects of Particulate Air Pollution on Ischemic Stroke and Hemorrhagic Stroke Mortality. Front Neurol. 2018;9. doi:10.3389/fneur.2018.00827

11. Cui M, Zhan C, Wu W, Guo D, Song Y. Acute Gaseous Air Pollution Exposure and Hospitalizations for Acute Ischemic Stroke: A Time-Series Analysis in Tianjin, China. Int J Environ Res Public Health. 2022;19(20):13344. doi:10.3390/ijerph192013344

12. Carod-Artal F. Introduction to tropical neurology. Epidemiology of tropical diseases. ResearchGate. Accessed May 2, 2025. https://www.researchgate.net/publication/286806322_Introduction_to_tropical_neurology_Epidemiology_of_tropical_diseases

13. Kang JH, Liu TC, Keller J, Lin HC. Asian dust storm events are associated with an acute increase in stroke hospitalisation. J Epidemiol Community Health. 2013;67(2):125–131. doi:10.1136/jech-2011-200794

14. Poulsen A. Air pollution and stroke; effect modification by sociodemographic and environmental factors. A cohort study from Denmark - ScienceDirect. June 2023. Accessed May 2, 2025. https://www.sciencedirect.com/science/article/abs/pii/S1438463923000561

15. Cunha MF, Pellino G. Environmental effects of surgical procedures and strategies for sustainable surgery. Nat Rev Gastroenterol Hepatol. 2023;20(6):399–410. doi:10.1038/s41575-022-00716-5

16. Zygourakis CC, Yoon S, Valencia V, et al. Operating room waste: disposable supply utilization in neurosurgical procedures. J Neurosurg. 2017;126(2):620–625. doi:10.3171/2016.2.JNS152442

17. Asamoto S, Sawada H, Muto J, Arai T, Kawamata T. Green Hospital as a new Standard in Japan: How far can Neurosurgery go in Japan? World Neurosurg. 2024;187:150–155. doi:10.1016/j.wneu.2024.04.086

18. Talibi SS, Scott T, Hussain RA. The Environmental Footprint of Neurosurgery Operations: An Assessment of Waste Streams and the Carbon Footprint. Int J Environ Res Public Health. 2022;19(10):5995. doi:10.3390/ijerph19105995

19. Zachem TJ, Chen SF, Venkatraman V, et al. Computer Vision for Increased Operative Efficiency via Identification of Instruments in the Neurosurgical Operating Room: A Proof-of-Concept Study. arXiv. Preprint posted online April 29, 2024. doi:10.48550/arXiv.2312.03001

20. Rigante L, Moudrous W, de Vries J, Grotenhuis AJ, Boogaarts HD. Operating room waste: disposable supply utilization in neurointerventional procedures. Acta Neurochir (Wien). 2017;159(12):2337–2340. doi:10.1007/s00701-017-3366-y

21. Shah S, George J, Barnard H. (PDF) The Carbon Footprint of Spinal Surgery: Experiences of a Tertiary Centre. September 2023. Accessed May 16, 2025. https://www.researchgate.net/publication/373661851_The_Carbon_Footprint_of_Spinal_Surgery_Experiences_of_a_Tertiary_Centre?cf_chl_tk=Nu8dO4EZLxbpR.ISutdqULHidoZaCqlWuXVzOu16ojI-1747400771-1.0.1.1-GdcAZKbZI9edZhjdf8dOeiJNGFbOVBfiBSi0zG3eAMk

22. Ross MN, Behrndt LW, McIntyre MK, Ross DA. Sustainability and Green Practices in the Neurosurgical Operating Room: A Scoping Literature Review. World Neurosurg. 2024;181:e752–e757. doi:10.1016/j.wneu.2023.10.123

23. Song J, Han K, Wang Y, et al. Microglial Activation and Oxidative Stress in PM2.5-Induced Neurodegenerative Disorders. Antioxidants. 2022;11(8):1482. doi:10.3390/antiox11081482

24. Cai M, Lin X, Wang X, et al. Long-term exposure to ambient fine particulate matter chemical composition and in-hospital case fatality among patients with stroke in China. Lancet Reg Health – West Pac. 2023;32. doi:10.1016/j.lanwpc.2022.100679

25. PM2.5 Exposure | State of Global Air. Accessed May 27, 2024. https://www.stateofglobalair.org/air/pm

26. Zhang W, Furtado K, Wu P, et al. Increasing precipitation variability on daily-to-multiyear time scales in a warmer world. Sci Adv. 2021;7(31):eabf8021. doi:10.1126/sciadv.abf8021

27. Alarcón R, Giménez B, Hernández AF, et al. Occupational exposure to pesticides as a potential risk factor for epilepsy. NeuroToxicology. 2023;96:166–173. doi:10.1016/j.neuro.2023.04.012

28. Guo Z, Ouyang W, Chen M, et al. Increasing precipitation deteriorates the progress of pesticide reduction policy in the vulnerable watershed. Npj Clean Water. 2023;6(1):1–10. doi:10.1038/s41545-023-00290-6

29. Caceres A, Caceres-Alan A, Caceres-Alan T. Toxoplasma gondii infections in pediatric neurosurgery. Childs Nerv Syst ChNS Off J Int Soc Pediatr Neurosurg. 2024;40(2):295–301. doi:10.1007/s00381-023-05915-2

30. Yan C, Liang LJ, Zheng KY, Zhu XQ. Impact of environmental factors on the emergence, transmission and distribution of Toxoplasma gondii. Parasit Vectors. 2016;9:137. doi:10.1186/s13071-016-1432-6

31. Chien LC, Sy F, Pérez A. Identifying high risk areas of Zika virus infection by meteorological factors in Colombia. BMC Infect Dis. 2019;19(1):888. doi:10.1186/s12879-019-4499-9

32. Fernandes S, Pinto M, Barros L, et al. The economic burden of congenital Zika Syndrome in Brazil: an overview at 5 years and 10 years. BMJ Glob Health. 2022;7(7):e008784. doi:10.1136/bmjgh-2022-008784

33. Mapping the global neurosurgery workforce. Part 1: Consultant neurosurgeon density in: Journal of Neurosurgery Volume 141 Issue 1 (2024) Journals. Accessed October 3, 2025. https://thejns.org/view/journals/j-neurosurg/141/1/article-p1.xml

34. Dewan MC, Baticulon RE, Rattani A, Johnston JM, Warf BC, Harkness W. Pediatric neurosurgical workforce, access to care, equipment and training needs worldwide. Neurosurg Focus. 2018;45(4):E13. doi:10.3171/2018.7.FOCUS18272

35. Kwakye G, Brat GA, Makary MA. Green surgical practices for health care. Arch Surg Chic Ill 1960. 2011;146(2):131-136. doi:10.1001/archsurg.2010.343

36. Mishra LD, Agarwal A, Singh AK, Sriganesh K. Paving the way to environment-friendly greener anesthesia. J Anaesthesiol Clin Pharmacol. 2024;40(1):9–14. doi:10.4103/joacp.joacp_283_22

37. Wang AY, Ahsan T, Kosarchuk JJ, Liu P, Riesenburger RI, Kryzanski J. Assessing the Environmental Carbon Footprint of Spinal versus General Anesthesia in Single-Level Transforaminal Lumbar Interbody Fusions. World Neurosurg. 2022;163:e199–e206. doi:10.1016/j.wneu.2022.03.095

38. Hoyler MM, White RS, Mack PF, Kelleher DC. Environmental report cards in anesthesia care: A quality metric for patients, providers and institutions. J Clin Anesth. 2021;73:110355. doi:10.1016/j.jclinane.2021.110355

39. Hammer S, Eichlseder M, Klivinyi C, et al. Effects of departmental green anaesthesia interventions on carbon dioxide equivalent emissions: a systematic review. Br J Anaesth. 2025;135(1):79–88. doi:10.1016/j.bja.2025.03.038

