## Supplemental Table 1 for "Assessing Global Neurosurgical Disease Burden Related To Climate Change: A Scoping Review"

**Supplementary Table 1:**

Search Terms*

| **Database** | **Search Terms** |
| --- | --- |
| PubMed | ("Neurosurgery"[Mesh] OR "Cerebrovascular Disorders"[Mesh] OR "Brain Injuries"[Mesh] OR "Epilepsy"[Mesh] OR "Brain neoplasms"[MeSH] OR neurosurg*[tiab] OR (neurolog* AND surg*) OR traumatic-brain-injur*[tiab] OR spinal-cord-injur*[tiab] OR spine-injur*[tiab] OR brain-tumor*[tiab] OR brain-tumour*[tiab] OR stroke*[tiab] OR hydrocephalus[tiab] OR epileps*[tiab] OR seizure*[tiab] OR brain-aneurysm*[tiab] OR disc-hernia*[tiab] OR degenerative-spinal-disease*[tiab] OR spina-bifida*[tiab] OR intracranial-abcess*[tiab] OR chiara-malformation* OR brain-cancer*[tiab] OR brain-neoplas*[tiab] OR brain-tumor*[tiab])  AND  ("Climate Change"[Mesh] OR "Environmental Pollutants"[Mesh] OR "Air pollution"[Mesh] OR "global warming"[MeSH] OR "Wildfires"[Mesh] OR "Floods"[Mesh] OR "Extreme Weather"[Mesh] OR "global warming"[tiab] OR "hot temperature*"[tiab] OR "Climatic Processes"[Mesh] OR greenhouse-effect*[tiab] OR "Carbon Footprint"[Mesh] OR high-temperature*[tiab] OR hot-environment*[tiab] OR heat-stress*[tiab] OR environmental-monitor*[tiab] OR extreme-temperature*[tiab] OR extreme-heat*[tiab] OR extreme-weather*[tiab] OR global-temperature*[tiab] OR heat-wave*[tiab] OR climate-change*[tiab] OR climate-data[tiab] OR ambient-temperature*[tiab] OR Severe-weather[tiab] OR Air-pollution[tiab] OR Wildfire*[tiab] OR Vector-borne-illness[tiab] OR vector-borne-disease*[tiab] OR Vector-ecology[tiab] OR Water-quality-impact*[tiab] OR Water-pollut*[tiab] OR Environmental-degradation[tiab] OR Rising-temperature*[tiab] OR CO2-Emission*[tiab] OR methane-emission*[tiab]) |
| Embase | ('neurosurgery'/exp OR 'cerebrovascular disease'/exp OR 'brain injury'/exp OR 'epilepsy'/exp OR 'brain tumor'/exp OR 'cerebrovascular disorder*':ti,ab OR 'cerebrovascular disease*':ti,ab OR neurosurg*:ti,ab OR 'neurological surg*':ti,ab OR (neurolog* AND surg*) OR 'traumatic brain injur*':ti,ab OR 'spinal cord injur*':ti,ab OR 'spine injur*':ti,ab OR 'brain tumour*':ti,ab OR stroke*:ti,ab OR hydrocephalus:ti,ab OR epileps*:ti,ab OR seizure*:ti,ab OR 'brain aneurysm*':ti,ab OR 'disc hernia*':ti,ab OR 'degenerative spinal disease*':ti,ab OR 'spina bifida*':ti,ab OR 'intracranial abcess*':ti,ab OR 'chiara malformation*' OR 'brain cancer*':ti,ab OR 'brain neoplas*':ti,ab OR 'brain tumor*':ti,ab)  AND  ('climate change'/exp OR 'greenhouse effect'/exp OR 'wildfire'/exp OR 'flooding'/exp OR 'extreme weather'/exp OR 'carbon footprint'/exp OR 'global warming':ti,ab OR 'hot temperature*':ti,ab OR 'greenhouse effect*':ti,ab OR 'high temperature*':ti,ab OR 'hot environment*':ti,ab OR 'heat stress*':ti,ab OR 'environmental monitor*':ti,ab OR 'extreme temperature*':ti,ab OR 'extreme heat*':ti,ab OR 'extreme weather*':ti,ab OR 'global temperature*':ti,ab OR 'heat wave*':ti,ab OR 'climate change*':ti,ab OR 'climate data':ti,ab OR 'ambient temperature*':ti,ab OR 'severe weather':ti,ab OR 'air pollution':ti,ab OR wildfire*:ti,ab OR 'vector borne illness':ti,ab OR 'vector borne disease*':ti,ab OR 'vector ecology':ti,ab OR 'water quality impact*':ti,ab OR 'water pollut*':ti,ab OR 'environmental degradation':ti,ab OR 'rising temperature*':ti,ab OR 'co2 emission*':ti,ab OR 'methane emission*':ti,ab OR 'climatic process*':ti,ab OR 'air pollution'/exp OR 'water pollution'/exp OR 'soil pollution'/exp OR 'salinization'/exp OR 'plastic pollution'/exp) |
| Scopus | ('neurosurgery'/exp OR 'cerebrovascular disease'/exp OR 'brain injury'/exp OR 'epilepsy'/exp OR 'brain tumor'/exp OR 'cerebrovascular disorder*':ti,ab OR 'cerebrovascular disease*':ti,ab OR neurosurg*:ti,ab OR 'neurological surg*':ti,ab OR (neurolog* AND surg*) OR 'traumatic brain injur*':ti,ab OR 'spinal cord injur*':ti,ab OR 'spine injur*':ti,ab OR 'brain tumour*':ti,ab OR stroke*:ti,ab OR hydrocephalus:ti,ab OR epileps*:ti,ab OR seizure*:ti,ab OR 'brain aneurysm*':ti,ab OR 'disc hernia*':ti,ab OR 'degenerative spinal disease*':ti,ab OR 'spina bifida*':ti,ab OR 'intracranial abcess*':ti,ab OR 'chiara malformation*' OR 'brain cancer*':ti,ab OR 'brain neoplas*':ti,ab OR 'brain tumor*':ti,ab)  AND  ('climate change'/exp OR 'greenhouse effect'/exp OR 'wildfire'/exp OR 'flooding'/exp OR 'extreme weather'/exp OR 'carbon footprint'/exp OR 'global warming':ti,ab OR 'hot temperature*':ti,ab OR 'greenhouse effect*':ti,ab OR 'high temperature*':ti,ab OR 'hot environment*':ti,ab OR 'heat stress*':ti,ab OR 'environmental monitor*':ti,ab OR 'extreme temperature*':ti,ab OR 'extreme heat*':ti,ab OR 'extreme weather*':ti,ab OR 'global temperature*':ti,ab OR 'heat wave*':ti,ab OR 'climate change*':ti,ab OR 'climate data':ti,ab OR 'ambient temperature*':ti,ab OR 'severe weather':ti,ab OR 'air pollution':ti,ab OR wildfire*:ti,ab OR 'vector borne illness':ti,ab OR 'vector borne disease*':ti,ab OR 'vector ecology':ti,ab OR 'water quality impact*':ti,ab OR 'water pollut*':ti,ab OR 'environmental degradation':ti,ab OR 'rising temperature*':ti,ab OR 'co2 emission*':ti,ab OR 'methane emission*':ti,ab OR 'climatic process*':ti,ab OR 'air pollution'/exp OR 'water pollution'/exp OR 'soil pollution'/exp OR 'salinization'/exp OR 'plastic pollution'/exp) |

*All databases searched on 6/27/24.
